# Intergenerational effects of parental educational attainment on parenting and childhood educational outcomes: Evidence from MoBa using within-family Mendelian randomization

**DOI:** 10.1101/2023.02.22.23285699

**Authors:** Alexandra Havdahl, Amanda M Hughes, Eleanor Sanderson, Helga Ask, Rosa Cheesman, Ted Reichborn-Kjennerud, Ole A. Andreassen, Elizabeth C. Corfield, Laurie Hannigan, Per Magnus, Pål R. Njølstad, Camilla Stoltenberg, Fartein Ask Torvik, Ragnhild Brandlistuen, George Davey Smith, Eivind Ystrom, Neil M Davies

**Affiliations:** Medical Research Council Integrative Epidemiology Unit at the University of Bristol, BS8 2BN, United Kingdom; Centre for Genetic Epidemiology and Mental Health, Norwegian Institute of Public Health, Oslo, Norway; Nic Waals Institute, Lovisenberg Diaconal Hospital, Oslo, Norway; Population Health Sciences, Bristol Medical School, University of Bristol, Barley House, Oakfield Grove, Bristol, BS8 2BN, United Kingdom; PROMENTA Research Center, Department of Psychology, University of Oslo, Oslo, Norway; Department of Child Health and Development, Norwegian Institute of Public Health, Oslo, Norway; NORMENT Centre, Division of Mental Health and Addiction, Oslo University Hospital & Institute of Clinical Medicine, University of Oslo, Oslo, Norway; Mohn Center for Diabetes Precision Medicine, Department of Clinical Science, University of Bergen, Bergen, Norway; Children and Youth Clinic, Haukeland University Hospital, Bergen, Norway; Centre for Fertility and Health, Norwegian Institute of Public Health, Oslo, Norway; K.G. Jebsen Center for Genetic Epidemiology, Department of Public Health and Nursing, Norwegian University of Science and Technology, Norway; Division of Psychiatry, University College London, Maple House, 149 Tottenham Court Rd, London W1T 7NF; Department of Statistical Sciences, University College London, London WC1E 6BT, UK

**Keywords:** MoBa, Mendelian randomization, educational attainment, indirect genetic effects, dynastic effects

## Abstract

The intergenerational transmission of educational attainment from parents to their children is one of the most important and studied relationships in social science. Longitudinal studies have found strong associations between parents’ and their children’s educational outcomes, which could be due to the effects of parents. Here we provide new evidence about whether parents’ educational attainment affects their parenting behaviours and children’s early educational outcomes using within-family Mendelian randomization and data from 40,879 genotyped parent-child trios from the Norwegian Mother, Father and Child Cohort (MoBa) study. We found evidence suggesting that parents’ educational attainment affects their children’s educational outcomes from age 5 to 14. More studies are needed to provide more samples of parent-child trios and assess the potential consequences of selection bias and grandparental effects.

## Introduction

Parents’ educational attainment correlates with various educational and non-educational outcomes in children. Differences in children’s educational outcomes emerge early in childhood and are highly persistent[1]. Exams taken in adolescence are highly predictive of final educational attainment in adulthood. The correlations between parents and their children could be due to direct genetic inheritance, effects of parents on their children via parenting behaviours, assortative mating, or demographic differences across the population[2]. The relative contribution of direct genetic inheritance and factors such as parenting is unclear[3]. Twin studies suggest that the heritability of educational attainment due to direct genetic effects is around 40%[4]. Genome-wide association studies (GWAS) suggest that the heritability of educational attainment due to common genetic variants is at least 11%[5]. However, there is increasing evidence that some of the associations identified in GWAS, particularly socioeconomic and psychological phenotypes, can be attributed to indirect genetic effects[3,5–7].

Most GWAS use samples of unrelated individuals in which it is challenging to control fully for indirect sources of association between genotypes and phenotypes, such as population structure or parental effects. Sibling designs provide an appealing approach for controlling for these factors because full biological siblings share their parents and, before conception, have an equal probability of inheriting any given variant from their parents[8,9]. However, a limitation of sibling designs is that these methods can only control for indirect genetic effects; they generally cannot be used to estimate the contributions of parents to their children’s outcomes. A complementary design uses data from mother, father and child ‘trios’. Within a family, and conditional on parental genotype, the inheritance of genetic variation from parents to children should be random with respect to the parents’ ancestry and parents’ assortment before conception. Thus, trio designs naturally control for many potential sources of bias seen in population-based GWAS. Previous studies have used trio designs to investigate the associations of polygenic indices and educational outcomes in the Norwegian Mother, Father and Child Cohort Study (MoBa).[10] However, it is possible to go further and estimate both the direct effects of genotype on the children and the effects of parents on their children via Mendelian randomization and instrumental variable analysis[2,11]. **Figure 1** illustrates these relationships.

**Figure 1:**
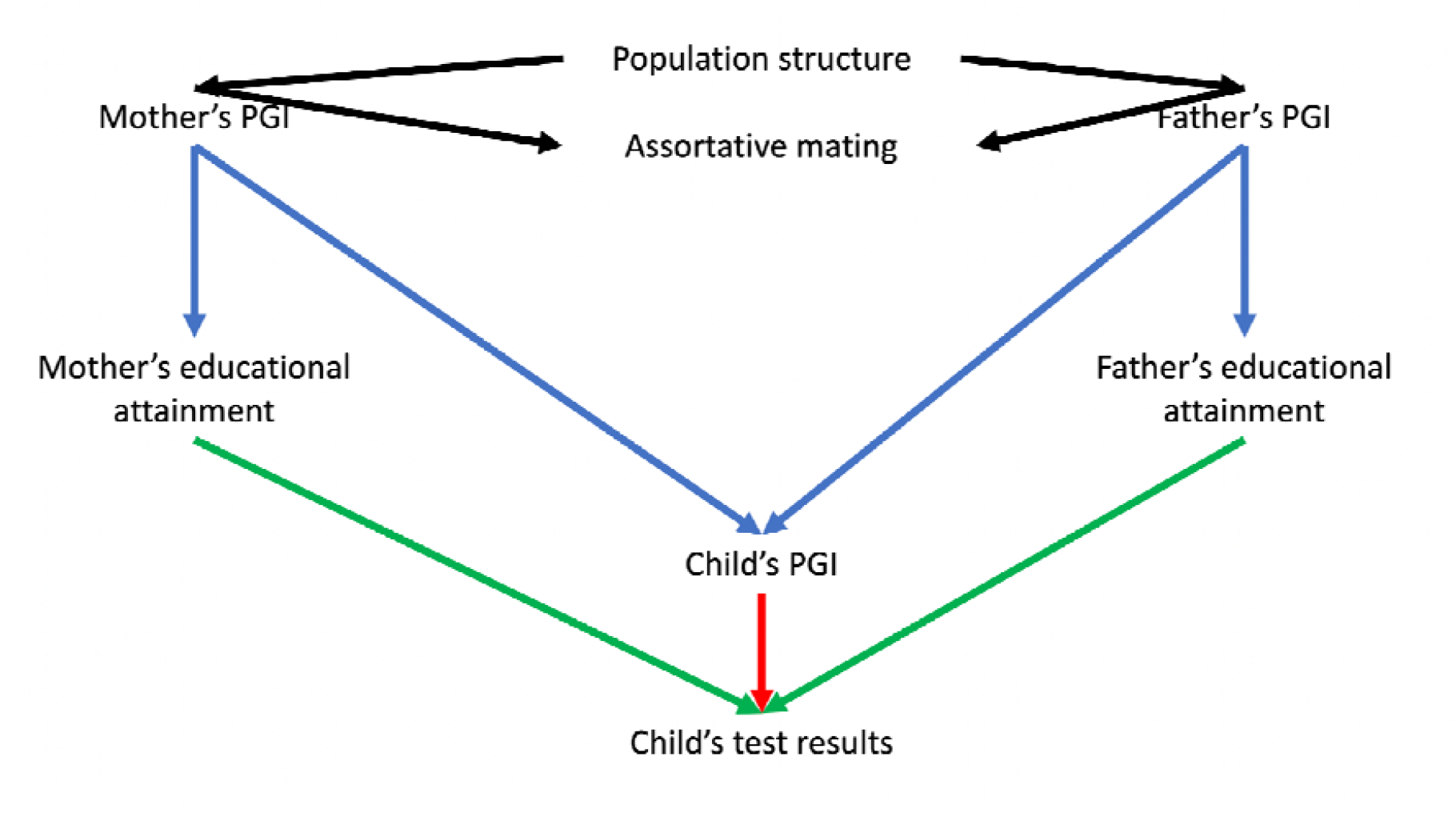
Directed acyclic graph illustrating the relationships between the mother and father’s polygenic indices (PGI), educational attainment and their child’s PGIs and test scores. Notes: The parental educational attainment polygenic index (PGI) can associate with children’s educational outcomes either via population structure, assortative mating (black arrows), dynastic effects (effects of parents on their children, including indirect genetic effects, green arrow), or direct transmission of education-associated variants (red arrow). We can precisely control for the direct transmission of DNA by adjusting for the PGI for educational attainment inherited by the child. The dynastic effects of parents’ educational attainment on their children’s outcomes can be estimated using multivariable Mendelian randomization, in which each parent’s educational attainment was instrumented by their index. We control for population structure using the principal components. Differential effects of mothers and fathers are unlikely to be explained by assortative mating, which will have symmetric biases on the estimated effects of mothers and fathers, see simulations for more details.

### Mendelian randomization

Mendelian randomization is an analysis method which uses genetic variants as instrumental variables to estimate the effects of the exposure of interest on the outcomes[12–14]. Instrumental variables are defined by three assumptions, i) “relevance”: they associate with the exposure of interest (parents’ educational attainment); ii) “independence”: there are no common causes of the instrument and the outcomes (children’s early educational outcomes and parenting); and iii) “exclusion”: the instruments only affect the outcomes via the trait of interest. Genetic variants are plausible instrumental variables because the transmission of genetic variants from parents to children is quasi-random. Germline genetic variation generally does not change across the life course, eliminating the risk of reverse causation[12,15]. The first assumption holds and is empirically testable; thousands of genetic variants robustly associate with educational attainment in a GWAS meta-analysis of three million individuals[16]. These genetic variants can be aggregated into polygenic indices. The second assumption could be violated if, as is likely, there are direct genetic effects (e.g. biological effects) of genetic variants associated with educational attainment on the children’s outcomes or dynastic effects (e.g. effects of parenting).

This paper uses multivariable Mendelian randomization to estimate the effects of the two exposures of interest-fathers’ and mothers’ educational attainment-on children’s outcomes and parenting behaviours (see **Methods** for details)[8,11,17]. The effect of each parent’s educational attainment can be identified by using their polygenic index for educational attainment as an instrument for their educational attainment. However, on average, children’s polygenic indices are 0.5 correlated with their parents’ polygenic indices. Thus, any direct effect of children’s polygenic indices on the outcomes would bias standard Mendelian randomization estimates of the effects of parents’ educational attainment. This bias can be controlled by including the children’s polygenic indices as covariates.

Parents’ polygenic indices for educational attainment could associate with their children’s educational outcomes for the following reasons that are not due to an effect of parents’ educational attainment on their children’s outcomes. First, these variants could systematically differ in frequency across the population (population structure). If these differences relate to differences in rates of educational attainment (e.g. between richer and poorer areas of a country), then this could confound the association of genetic variants and educational attainment. Second assortative mating, as more educated mothers are likely to have more educated spouses. Third, other family members, such as grandparents, whose genotypes correlate with the parents, could induce associations between parents’ genotypes and their children’s outcomes. Here we control population structure using principal components of genetic variation[18]. We assess the impact of assortative mating via directed acyclic graphs and simulations.

Here we provide new evidence about the familial and social mechanisms that mediate the associations between parents’ and children’s genetic variation and children’s educational outcomes. We estimated the effects of parents’ educational attainment on their children’s academic skills and support using a sample of births from the MoBa study. This sample was linked to nationally standardised test data for students between nine and fourteen years old and questionnaire measures of parenting behaviour for children aged five and eight.

## Results

After applying our inclusion criteria, 40,879 complete mother-father-child trios were eligible for analysis. See **Supplementary Figure 1** for a STROBE flow diagram of the inclusion and exclusion from the analysis sample[19]. **Supplementary Table 1** shows the characteristics of the participants for the raw and imputed data. The participating children were 51% male, and their mothers and fathers, on average, had 17.3 and 16.5 years of educational attainment, respectively. This compares to 15.2 years in the adult Norwegian population in 2007.[20] There was little evidence that the samples differed between participants, with no and any missing values of exposures or outcomes. Below we present the results estimated using multiple imputation; please see the supplementary information for non-imputed results (**Supplementary Tables 2, 3 and 4**).

### Association of polygenic indices and parental educational attainment

The child polygenic index, constructed from 1,729 independent genome-wide significant variants from Okbay and colleagues[16], was associated as expected with the mother and father polygenic indices (linear regression coefficients 0.498 (95% confidence interval (95%CI) 0.492 to 0.505) and 0.496 (95%CI 0.489 to 0.502), respectively). Fathers’ and mothers’, but not children’s, educational attainment polygenic indices were associated with the mothers’ and fathers’ educational attainment (**Supplementary Table 5**). In the multivariable-adjusted analysis, trios with one standard deviation higher child, mother and father polygenic indices had a mother with 0.01 (95%CI -0.02 to 0.04), 0.54 (95%CI: 0.51 to 0.57) and 0.31 (95%CI 0.28 to 0.34) additional years of education respectively. Equivalent increases in the father’s years of education were 0.03 (95%CI -0.01 to 0.06), 0.40 (95%CI 0.37 to 0.43), and 0.62 (95%CI 0.59 to 0.65) additional years, respectively. The mother and fathers’ polygenic indices strongly predicted the parents’ educational attainment. Conditional on the children’s and fathers’ polygenic index, the mothers’ index explained 3.85% of the variation in mothers’ educational attainment with a conditional F-statistic of 588.4. The equivalent figures for fathers were 3.84% and 579.6, respectively[21]. Six hundred twenty-one mothers from MoBa were included in the educational attainment GWAS, which is unlikely to bias our results[22].

### Phenotypic associations of parents’ education and child test scores

Mothers’ and fathers’ educational attainment were similarly phenotypically associated with higher test scores for the children across all tests and all ages (**Figure 2**). For example, a one-year increase in mothers’ educational attainment was phenotypically associated with an 0.04 (95%CI: 0.04 to 0.05) SD increase in the same test. Fathers’ educational attainment was phenotypically associated with an 0.05 (95%CI: 0.05 to 0.06) standard deviation (SD) increase in the child’s English test scores in grade 5.

**Figure 2:**
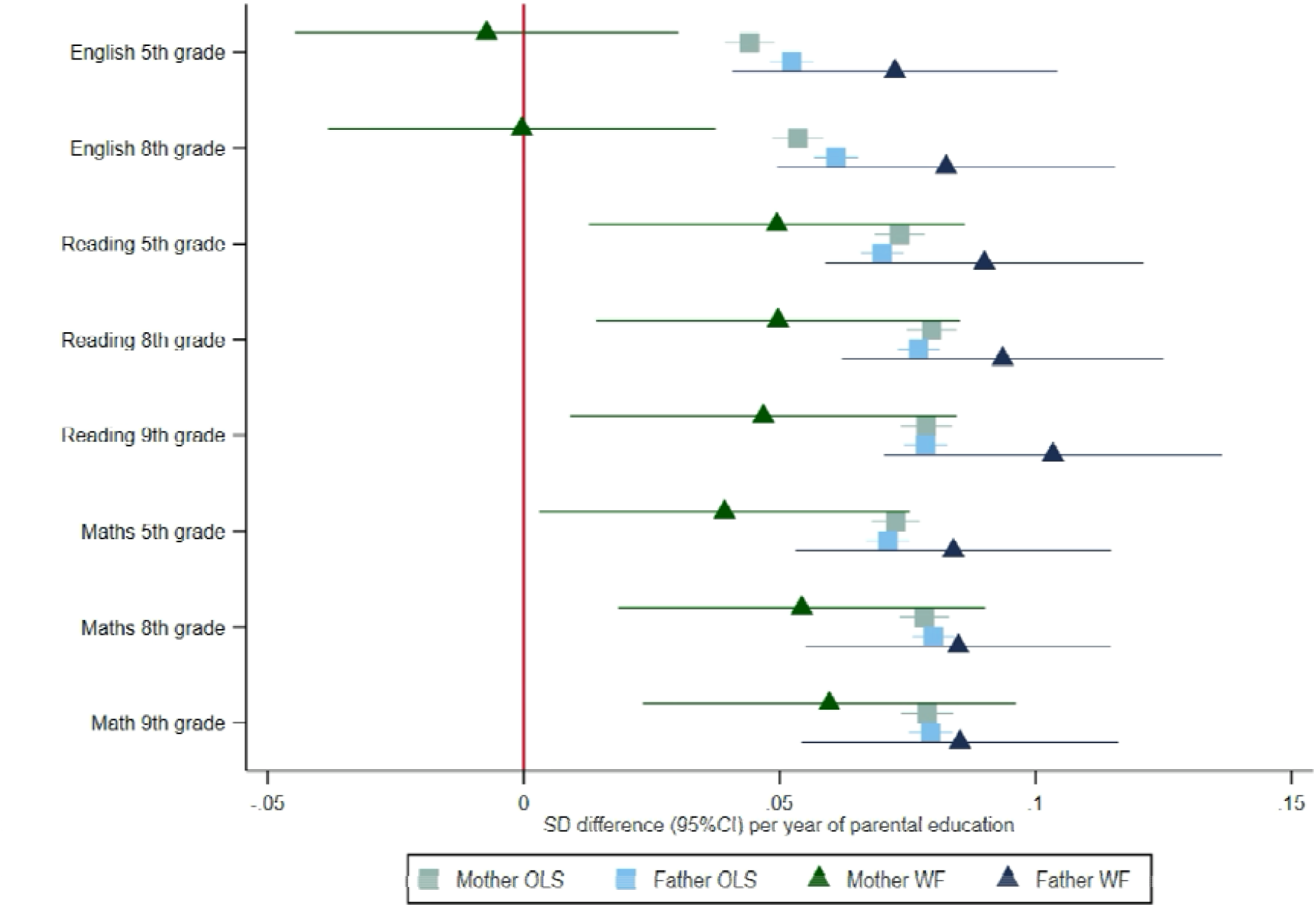
Effect of parents’ educational attainment on children’s nationally standardised test scores, estimated using multivariable-adjusted regression (OLS) and within-family Mendelian randomization (WF), estimated on the full sample using multiple imputation (N=40,879). Mean difference in standard deviations and 95% confidence intervals in test scores per year of parental education reported.

### Within-family Mendelian randomization estimates of the effect of parents’ education on child test scores

The within-family Mendelian randomization estimates largely provided consistent evidence that mothers’ and fathers’ educational attainment increased their children’s test scores. Two exceptions were that we found little evidence that mothers’ educational attainment affected English test results in grades 5 and 8. The Mendelian randomization estimates of the effects of mothers’ educational attainment on reading and maths test scores were slightly smaller than, but consistent with the phenotypic associations for reading and maths in grades 5, 8 and 9. The within-family Mendelian randomization estimates of fathers’ educational attainment effects were consistent with the phenotypic associations (**Figure 2**).

### Parenting behaviours and school readiness age five

We investigated if there was any evidence that parental educational attainment affected parenting behaviours or school readiness using the mothers’ responses to questionnaires at ages five and eight. **Figure 3** and **Supplementary Table 5** present phenotypic and within-family Mendelian randomization estimates of these relationships. Mothers’ and fathers’ educational attainment was phenotypically associated with slightly lower levels of reporting they taught their child letters, higher levels of reporting that their child enjoyed being read to, and literacy scores. In addition, there was evidence that mothers’, but not fathers’, years of education were phenotypically associated with higher communication narrative scores. Overall, the phenotypic associations were small. The within-family Mendelian randomization estimates were less precise and generally consistent with the null and the phenotypic associations.

**Figure 3:**
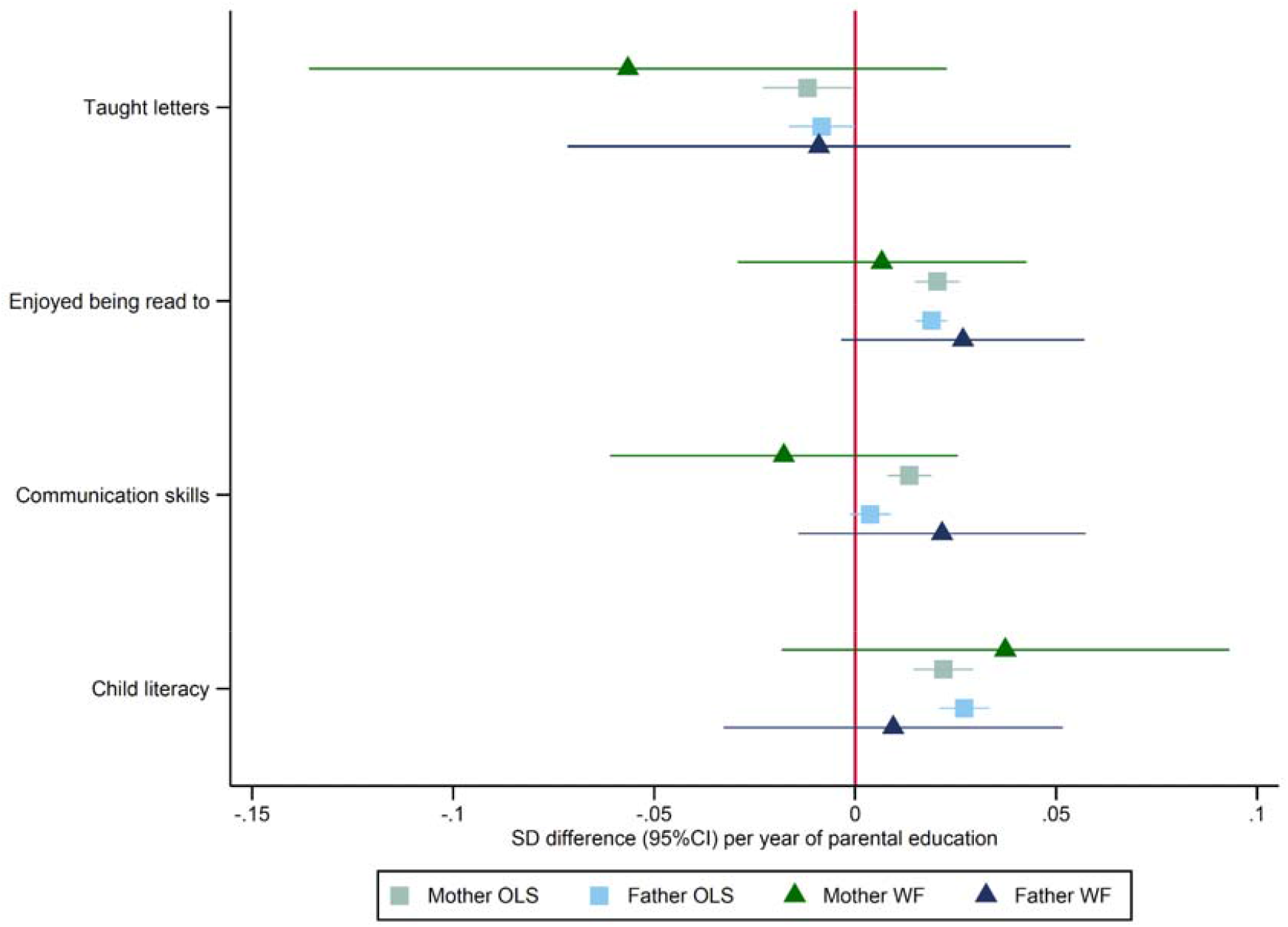
Effect of parents’ educational attainment on questionnaire measures of reading and communication skills and parental nurturing at age 5, estimated using multivariable-adjusted regression (OLS) and within-family Mendelian randomization (WF), estimated on the full sample using multiple imputation (N=40,879).

### Parenting behaviours and early educational measures at age eight

Both mothers’ and fathers’ educational attainment were phenotypically associated with questionnaire measures of parenting and early educational outcomes reported by the mother (**Figure 4**). Children of more educated mothers and fathers had higher teacher assessments and language skills and reading behaviour scores, but lower reading mastery scores. Mothers’ and fathers’ education was phenotypically associated with reporting fewer hours of homework and help with homework at home and after school. The within-family Mendelian randomization estimates suggested that each additional year of mothers’ education decreased the number of hours of help with homework (-0.04, 95%CI: -0.07 to -0.01). The within-family Mendelian randomization estimates suggested that an additional year of fathers’ education increased the reading score by 0.16 (95%CI: 0.08 to 0.24) units.

**Figure 4:**
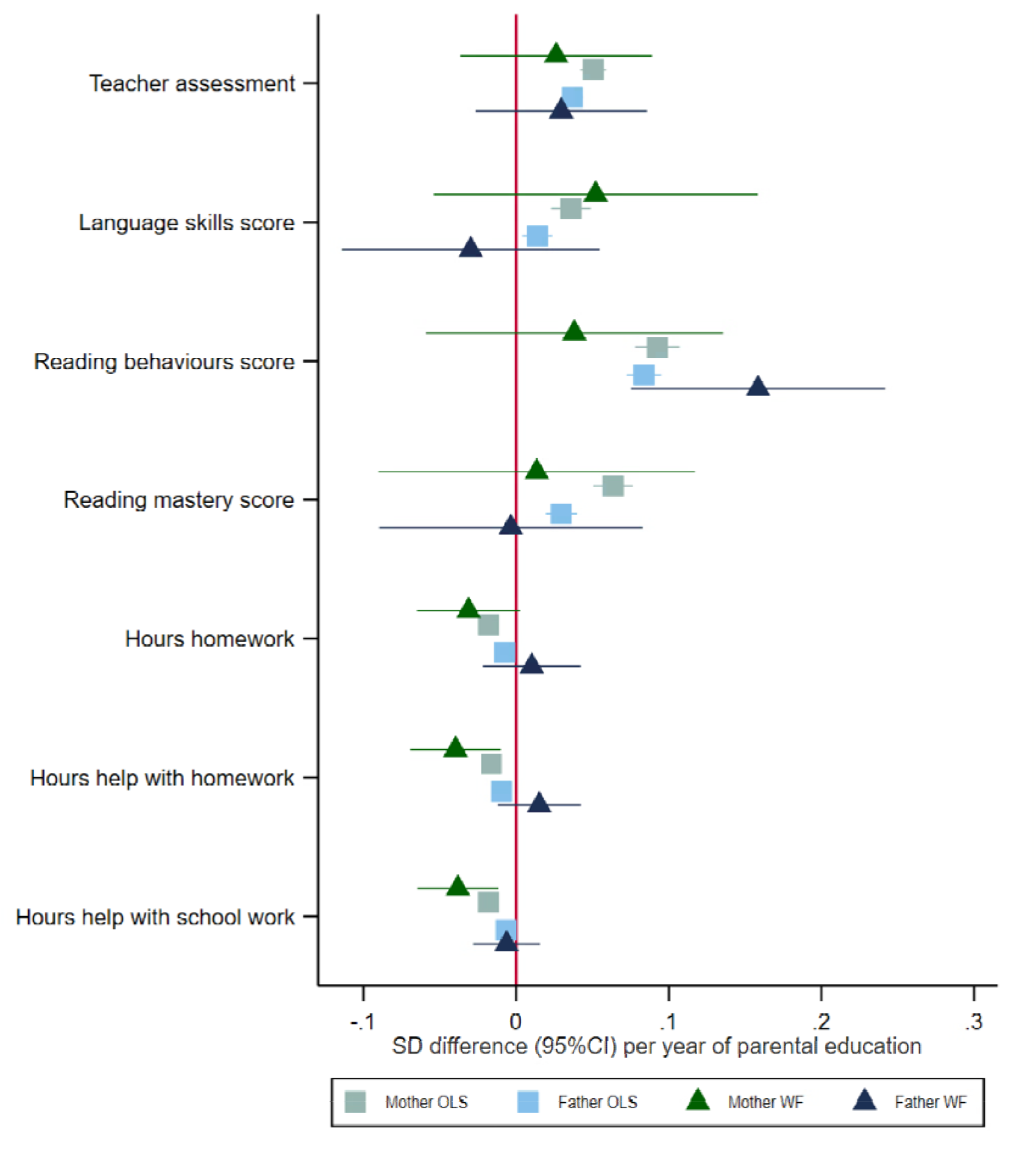
Effect of parents’ educational attainment on questionnaire measures of early educational attainment and parental nurturing at age 8, estimated using multivariable-adjusted regression (OLS) and within-family Mendelian randomization (WF), estimated on the full sample using multiple imputation (N=40,879).

At age 8, most of the estimates were similar between males and females. We detected an effect of fathers’ educational attainment on reading behaviours for females and males. We detected an effect of mothers’ educational attainment on fewer hours of help with homework at home and school for males. There was little evidence that these estimates systematically differed between male and female children. The stratified results were less precise than the pooled results because of the reduced sample size.

#### Non-imputed data

We repeated the analysis using non-imputed data. The sample size dropped substantially (min N=15,420, max N=39,163), particularly for the questionnaire measures, and consequently, non-imputed estimates were less precise. However, the non-imputed results were consistent with the primary analyses.

#### SNP level two-sample Mendelian randomization

One explanation for our results is that the variants have horizontally pleiotropic effects in the parents, and are due to other related factors such as cognition. We investigated this by estimating the associations of each parental SNP and each outcome adjusting for the child’s SNP. We used summary data (two-sample) Mendelian randomization methods to estimate the effects of parental attainment on each outcome. We used the inverse variance weighted estimator (IVW) (which assumes no directional pleiotropy), the weighted median (which assumes that at least 50% of the weighting comes from SNPs that have no pleiotropic effects), and the weighted mode estimator (which assumes that a plurality of SNPs have no pleiotropic effects), and MR-Egger (which assumes the pleiotropic effect is uncorrelated with the magnitude of the SNP-exposure association). Supplementary Table S8 reports these results. The IVW estimates were more precise than, but largely consistent with the single-sample estimates. The IVW results detected effects of mothers’ education on English test results in grade 5 and 8, and of both mothers’ and fathers’ education on some of the questionnaire measures at ages 5 and 8. The MR-Egger intercept generally provided little consistent evidence of pleiotropy or heterogeneity. Exceptions to this were the effects of mothers’ education on Maths test results in grade 5, 8, and 9 and enjoying being read to at age 5.

Differences in the effects of education on household income for men and women could explain our results. We investigated if years of educational attainment of mothers had differential effects on household income than fathers. However, the implied effects of an additional year of education were almost identical. The estimates implied that each additional year of schooling increased household income by 23,055 (95%CI: 15,397 to 30,713) and 22,535 (95%CI: 16012 to 29058) Norwegian Krone for men and women respectively ($2144 and $2096 at 2023 exchange rates). Thus, mothers’ and fathers’ educational attainment had similar effects on household income, which is unlikely to explain our results.”

### Simulations

We investigated whether our results could be explained by assortative mating using simulations. Both ordinary least squares and standard Mendelian randomization estimates of the effects of parents on their children were biased (**Supplementary Figure 8**). However, we found that assortative mating did not bias within-family Mendelian randomization analyses when both parents are included in a multivariable estimation. By including both parents in the estimation correlation between their genetic scores and phenotypes are adjusted for, removing any bias from assortative mating. See methods for details of the simulations.

## Discussion

We investigated the associations of polygenic indices for educational attainment and nationally standardised test scores in school and questionnaire measures of parenting behaviour, school readiness and early educational outcomes. Both mothers and fathers’ educational attainment strongly predicted children’s test scores in schools. Our within family Mendelian randomization results suggest that mothers’ and fathers’ educational attainment affected reading and Maths test results after controlling for direct genetic effects. The within family Mendelian randomization results from the questionnaire responses on parenting behaviour, school readiness and early educational outcomes were less precise, but suggested that fathers’ educational attainment increased reading behaviours, and mothers’ educational attainment resulted in their children receiving fewer hours help with academic work. Our results were consistent when we stratified by sex, used the non-imputed data, and used two-sample summary data Mendelian randomization.

### The context within the literature

The intergenerational transmission of traits, including educational attainment, has been the focus of an extraordinary amount of research across many fields[23–31]. Kong and colleagues (2018) found evidence of indirect genetic effects in an Icelandic study from deCODE[6]. They found that non-transmitted parental polygenic indices for educational attainment were 30% as strongly associated with the children’s educational outcomes as the children’s polygenic index. These results suggested that some aspect of the familial or social environment affected the children’s outcomes and that the association of the educational attainment polygenic index and education and other outcomes were unlikely to be solely driven by individual-level direct effects in the child. Consistent with this, Howe and colleagues conducted a within family sibling GWAS and found that the associations of SNPs with socioeconomic traits substantially attenuated within families.[9] Okbay and colleagues report similar results[16]. Again, these results and those from similar studies[5] suggest the SNP-educational attainment associations are partially due to indirect genetic effects in addition to the direct effects. While these results provided fascinating insights into how these phenotypes transmit from generation to generation, they have raised important new questions about which aspects of the familial or social environment mediate these associations.

Nivard and colleagues (2022) used a smaller, earlier release of the MoBa data (25,215 children) to investigate the relationship between non-cognitive and cognitive components of the educational attainment polygenic index[10]. They report comparable mother and father polygenic indices associations with children’s outcomes. Cheesman and colleagues (2022) used relatedness disequilibrium regression (RDR) to estimate the effects of genetic nurture on child anxiety and depression symptoms at age 8 using a sub-sample of MoBa[32]. A sibling-based model using 3,500 sibling pairs in the parents’ generation found little evidence that either parent’s polygenic indices indirectly affect children’s outcomes. However, these estimates had limited power to detect indirect genetic effects. Similarly, Isungset and colleagues (2022) use a sub-sample of the MoBa study and report similar associations of parents’ and children’s polygenic indices and educational outcomes[33]. Wertz and colleagues (2020) report findings from 860 mother-child pairs in the E-Risk study. They found that controlling for children’s polygenic index slightly attenuates the association of mothers’ polygenic index and child outcomes and associations between the mothers’ index and measures of nurturing behaviour[34]. Here we add to the literature by using Mendelian randomization to estimate the effects of parenting, while controling for assortative mating, direct genetic transmission, and reporting sensitivity analyses to investigate whether these associations could be due to horizontal pleiotropy.

Our findings are also consistent with non-genetic research, which has found that grandparents’ social class is associated with grandchildren’s outcomes, even after accounting for parents’ social class, income and wealth[35]. However, measurement error virtually guarantees that it will not be possible to fully control for parents’ socioeconomic position fully[36]. Studies using natural experiments of school reforms have found compelling evidence that the association between parents’ educational attainment and their children’s educational outcomes is likely to be partially causal[37]. Our findings are consistent with a registry-based non-genetic study in Norway[38]. This study found that the association of fathers’ educational attainment and their children’s outcomes attenuated if they died before the child left home. Similarly, an Israeli study used data from parental deaths, divorces and other family events and found evidence that parents’ educational attainment was more strongly associated with children’s outcomes when parents spent more time with their children[39].

A key innovation of this study is to provide new evidence about possible familial and social mechanisms that mediate the associations of SNPs and educational outcomes in children. If differences in the familial environment drive the attenuation of associations seen in within-family studies, we might expect associations between parental genetic variation and differences in nurturing behaviour. In this study, we directly tested this hypothesis using measures of directly reported behaviour. For many of the questionnaire measures, the Mendelian randomization estimates of the effects of parents’ educational attainment on the outcomes were consistent with the null and the phenotypic associations. There are two explanations for this, first a lack of statistical power and second that there is no effect of parental educational attainment on these aspects of parenting. Our Mendelian randomization results, while less precise than the phenotypic associations, are relatively precise and were able to exclude effects bigger than 0.1 of SD for most outcomes. Frequently the phenotypic associations were smaller than this, suggesting that if parents’ educational attainment does affect these aspects of education or parenting, their individual impacts are likely to be small.

### Strengths and limitations

A key strength of our study is that we used one of the largest samples of mother, father and child trios. The inheritance of genetic variants from parents to their children is quasi-random[40], and controlling for children’s polygenic indices allows us to control for direct genetic effects. Furthermore, MoBa has detailed questionnaire data on parenting behaviours and the parents’ reports about their children. Finally, we used Mendelian randomization to estimate the size of the effects of mothers’ and fathers’ educational attainment on each of the outcomes. A limitation of our results is that they may not be generalizable to other populations or familial relationships.

Our results could be due to the horizontal pleiotropic effects of the education-associated variants on closely related phenotypes in the parents (e.g. cognition). Genetic variants associated with educational attainment also associate with other phenotypes, such as cognition. Estimates from a range of pleiotropy robust two-sample Mendelian randomization estimators were consistent with our primary single-sample analysis. However, the pleiotropic effects of a closely related trait, such as cognition, potentially violate the assumptions of these methods. Future papers could investigate whether it is possible to combine multivariable Mendelian randomization and within-family study designs to test whether these findings are due to parents’ educational attainment or cognition. Furthermore, we must interpret Mendelian randomization estimates of categorical exposures like educational attainment with care. The genetic variants associated with educational choices across the life course may influence attainment differently in different periods or educational stages[41,42]. Future studies could use time-varying Mendelian randomization to investigate whether there is any heterogeneity in the effect of parental education by stage of educational attainment (e.g. remaining in school to age 18 versus attending university).

A key strength of MoBa is that it is a very large sample, single sampling frame, and excellent measurement of phenotypes, including high coverage nationally and standardised test indices measured on a continuous scale. In general, this helped maximise the statistical power of our study. However, we had relatively few measures of parenting behaviours, and these measures may not be the key mediating mechanisms. Furthermore, we did not investigate all aspects and features of parenting or genetic nurture or personality, such as openness or curiosity, and this may explain why we found relatively little evidence that our measures of parenting mediated the indirect genetic effects in our single sample analysis. Furthermore, despite our large sample, our study still had limited statistical power, which meant that we could not detect differences between the null, the phenotypic associations and the Mendelian randomization estimates for many outcomes. Future studies should use larger samples to investigate this hypothesis.

Within-family Mendelian randomization methods account for family-level factors when looking at the effects of the child’s risk factors. Here, however, we are estimating the effects of parent-level factors. We controlled for the children’s genotype, excluding the direct genetic effects of inheriting specific variants. However, other indirect genetic effects, aside from dynastic effects from parents, could explain our findings, for example, assortative mating on educational attainment in the parent’s generation. However, our simulations suggest that while assortative mating could explain bias in standard Mendelian randomization estimates, it is not sufficient to induce bias in within-family estimates that adjust for child genotype. Our models controlled for principal components of population stratification, so residual population structure is unlikely to explain our results. Another mechanism could be assortative mating and dynastic effects from the grandparent generation or via sibling effects[43]. Future studies should use larger samples of multi-generation families or imputed grandparental genotypes to investigate this hypothesis.

Finally, selection and collider bias could induce associations between genetic variants in parents and outcomes in their children if parental education and child test scores are related to the likelihood of being included in the analysis sample. More educated people are more likely to participate in studies like MoBa, and thus the complete trios, which sample and genotype the mother, father and their child, are likely to be a non-random sample from the population. This selection may be more strongly related to fathers than mothers, as fewer fathers participated and gave a DNA sample.

### Directions for future research

Future studies should recruit and analyse larger samples of parent-child trios and investigate more detailed and precise measurements of parenting behaviour in large representative cohorts of trios. Furthermore, the direct and indirect genetic effects may change after the children leave home. For example, at later stages of education, the importance of parental education may become more pronounced (e.g. navigating higher education choices). Future studies should follow up on the MoBa cohort of children once their educational attainment is complete. In addition, further study into the impact of selection bias on within-family methods could help elucidate whether this source of bias is likely to explain our results. Finally, future studies should explicitly model and estimate assortative mating and dynastic effects, e.g. using structural equation modelling and OpenMx, and techniques such as almost exact Mendelian Randomization[44,45].

## Conclusion

Parents’ educational attainment, or a closely related trait, is likely to affect their children’s test scores during school. In addition, the effects of fathers’ educational attainment appear to be greater than mothers, and there is some evidence that these effects may indicate differences in parenting behaviour. This study adds to the growing evidence of the importance of familial effects on children’s early educational outcomes.

## Methods

### Study population

The Norwegian Mother, Father and Child Cohort Study (MoBa) is a population-based pregnancy cohort conducted by the Norwegian Institute of Public Health. Participants were recruited from all over Norway from 1999-2008. The women consented to participation in 41% of the pregnancies. The cohort includes approximately 114,500 children, 95,200 mothers and 75,200 fathers.[46]. Blood samples were obtained from both parents during pregnancy and mothers and children (umbilical cord) at birth.[47] The current study is based on version 12 of the quality-assured questionnaire data files released for research in January 2019. The establishment of MoBa and initial data collection was based on a license from the Norwegian Data Protection Agency and approval from The Regional Committees for Medical and Health Research Ethics. The MoBa cohort is currently regulated by the Norwegian Health Registry Act. The current study was pre-approved by The Regional Committees for Medical and Health Research Ethics (2016/1702).

### Phenotypic measures

#### Exposures

##### Parents’ education

We defined parents’ educational attainment using data from the linked administrative data from Statistics Norway. We also used data from the mothers’ responses to questionnaires about their highest level of educational attainment. The questionnaire responses were included in the imputation model and used to impute a small number of missing values of the administrative data. We converted the parents’ responses to years of education as indicated by the International Standard Classification of Education (ISCED) categories for education. This mapping is shown in **Supplementary Table 6**.

#### Outcomes

##### Children’s test scores from grades 5, 8, and 9

We extracted national test scores from the linked administrative data from tests administered by the Norwegian Ministry of Education and provided by Statistics Norway.[48] These tests are nationally standardised and have been taken across schools in Norway since 2004. The data contain eight measures of educational achievement; for English in grades 5 and 8 and reading and Maths for grades 5, 8 and 9. These tests were graded on an ordinal scale of between 34 and 58 values. We normalised each test variable by year to have a mean of zero and a standard deviation of one. These administrative datasets were relatively complete; most participants (96%) had test score data. The tests are mandatory, but the schools can apply for exceptions for students with special education due to disabilities or a minority language.

##### Parenting behaviours and children’s school readiness at age 5

When MoBa children were five years old, the mothers answered a series of questions about parenting behaviour and their child’s school readiness. 1) Teaching letters: The mothers reported how frequently they taught how to a) print and b) read letters in a week on a five-item scale (“never”, “seldom”, “sometimes”, “often”, “very often”). These responses were combined to give a 0 to 10 scale. 2) Enjoys being read to: Mothers reported how long their child enjoyed being read to, choosing from “not read to”, “does not like being read to”, “< 5 minutes”, “6-15 minutes”, “16-45 minutes”, “more than 45 minutes”. This was treated as a 5-point scale, with not read to and does not like being read to both coded as zero. 3) Communication skills: Mothers also reported their child’s communication skills by rating their child’s ability to a) tell a story and b) communicate their needs on a three-item scale: 1=“very poor/poor”, 2=“average”, 3= “very good/good”. These responses were added together to result in a scale of 0 to 6. 4) Child literacy: Mothers answered five “yes” or “no” questions on whether the child was a) interested in writing letters, b) interested in books, c) able to read simple words, d) able to read simple sentences, and e) able to write their name. These responses were aggregated into a score of zero to five for the number of yes responses.

##### Education-related phenotypes age 8

At age 8, the mothers answered a series of questions about their child’s education. 1) <u>Teacher assessment</u>: Mothers reported the teacher’s assessment of their child’s reading skills in the first grade and their reading and arithmetic skills in the second grade. For each, mothers could describe the teacher’s assessment as either 1=“mastering well”, 2=“must work more, but teacher not concerned”, or 3=“teacher concerned”. Responses indicating do not know or not discussed with the teacher were coded as missing. These items were summed and reverse-coded to create a teacher assement score. 2) <u>Language skills score</u>: Finally, the mothers reported their child’s pronunciation-including how easy they were to understand, how easily strangers understood them, their child’s ability to tell a story, and how well they communicated their needs to others. We summed these items into a score. 3) <u>Reading behaviours score</u>: The mothers were asked how often they read to their child, how long their child liked to be read to for, how long they liked to read by themselves, and the type of books the child liked to read. These four responses were summed into a score. Frequency was measured on a Likert scale” 1=“never”, 2=“1-2 times a week”, 3=“3-4 times a week”, 4=“5-6 times a week”, and 5=“everyday”. Length of reading was indicated using the following Likert scale: 0=“does not like”, 1=“5 minutes or less”, 2=“6-15 minutes”, 3=“6-15 minutes”, 4=“16-25 minutes”, 5=“more than 45 minutes”, and 6=“never read to”. The last category was recoded to zero. 4) <u>Reading mastery score</u>: The mothers were asked whether their child had mastered reading via “yes”/”no” questions asking whether they could read simple stories out loud, identify letters, read suitable texts, write at least three sentences, and write a page of text. The responses were summed into a score and rescaled so that higher values indicate higher reading skills. A minority of the participants had missing values to these questions but were given an alternative set of questions. We imputed the values from the secondary questions using multiple imputation (see below for details of the imputation). 5) <u>Hours of homework</u>: The mother reported how many hours the children spent doing homework at home. 6) <u>Hours help with homework</u>: The number of hours help with homework at home. 7) <u>Hours help with schoolwork</u>: Hours help with school work at school or after school. The hours help with homework and schoolwork questions had the following possible responses: 1=“no homework”, 2=“0 hours”, 3=“1-2 hours”, 4=“3-4 hours”, 5=“5-6 hours”, and 6=“7 hours or more”. All variables were rescaled so that larger values indicated a more skilled outcome, and the smallest value was zero.

#### Covariates

We derived each child’s sex and year of birth from the Medical Birth Registry (MBRN), which is a national health registry containing information about all births in Norway. For other variables we imputed missing values using multiple imputations, including responses to the Q1 survey completed around the 15th week of pregnancy. In the imputation models, we also included measures of mothers’ and fathers’ educational attainment as reported by mothers and fathers in questionnaires completed around the 15th week of pregnancy. We coded educational attainment using the International Standard Classification of Education (ISCED) (**Supplementary Table 1**). All models control for child sex and year of birth, and the 20 principal components, genotyping centre and chip for mother, father and child.

### Genotyping, quality control and imputation

Genotyping of MoBa was conducted over several years by three research projects, with varying selection criteria, genotyping arrays, and genotyping centres. A standardised pipeline was used for pre-imputation quality control (QC), phasing, imputation, and post-imputation QC of the genotyping data. Full details of the MoBa genotyping efforts and the MoBaPsychGen QC pipeline are described elsewhere[49–51]. In this study we used the sub-sample of MoBa samples with genetic data that passed the MoBaPsychGen post-imputation QC (n = 76,577 children, 53,358 fathers, and 77,634 mothers).

#### Polygenic index for educational attainment

For each participant, we constructed a polygenic index for educational attainment. First, we selected variants to include in the index using summary data from Okbay and colleagues (2022) and 23andMe. This included a small sample of MoBa parents. Next, we selected all variants included in the GWAS, and the QC’d MoBa data that exceeded genome-wide significance levels (p < 5 × 10^-8^) and clumped to independent variants within 10,000 kb and r^2^ < 0.01. This process resulted in 1,729 SNPs, which we used to construct polygenic indices weighted by effect size for the children, mothers and fathers using the individual participant data.

### Statistical analysis

#### Statistical models

##### Individual-level data analysis

We estimated the effects of mothers’ and fathers’ educational attainment using three estimators. First, we estimated mutually-adjusted associations between mothers’ and fathers’ educational attainment and the outcomes using phenotypic multivariable-adjusted linear regression.

Second, we used individual participant data, a multivariable Mendelian randomization estimator. This applies a standard two-stage least squares instrumental variable estimator with two exposures and two instruments, the mother and fathers’ polygenic indices. These are valid instruments, conditional on including the child’s polygenic index as a covariate. We used the following model:

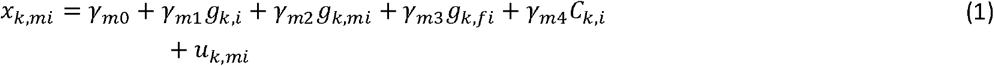

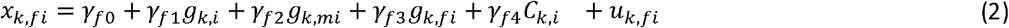

and

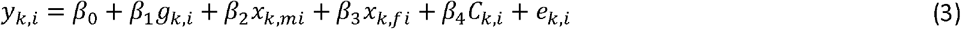

where, *y*_*k,i*_ and *x*_*k,mi*_ and *x*_*k,fi*_ are the outcome and mothers and fathers’ educational attainment for individual i from family *k*. The polygenic indices for the child, mother and father are *g*_*k,i*_, *g*_*k,mi*_ and *g*_*k,fi*_. The error terms are given by *u*_*k,i*_ and *e*_*k,i*_, and standard errors were robust and clustered by family *k*.

For comparison, as a sensitivity analysis, we report a conventional Mendelian randomization analysis, which does not control for the children’s polygenic indices. However, estimates of the effects of parents in this analysis will be biased by any direct effects of the child’s polygenic index on the outcomes. All analyses included the child, mother, and father’s genotyping centre, genotyping chip, child sex and year of birth, mother and father age, number of prior pregnancies, and the first 20 genetic principal components from the child as covariates.

##### Two-sample summary data Mendelian randomization

Third, we investigated whether our results were due to horizontal pleiotropy by applying two-sample summary data Mendelian randomization estimators.[13,52] These methods use SNP-level data for SNPs included in polygenic indices, comparing the associations of individual genetic variants and the exposures with the same variants’ associations with the outcomes. For this analysis, we clumped the GWAS summary data to 510 independent variants within 10,000 kb and r^2^ < 0.001. We used a more conservative threshold because in contrast to single sample estimators using polygenic indices, two sample estimators do not by default account for the correlation between SNPs included in the analysis. We estimated SNP-level associations with outcomes using linear models and individual-level imputed data, which mutually adjusted for mother, father and children’s genotypes and clustered standard errors within related families. We used the following model:

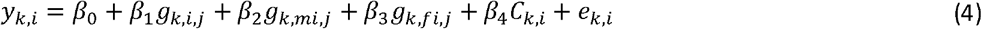

Where *g*_*k,i,j*_, *g*_*k,mi,j*_, and *g*_*k,fi,j*_ are the genotype at locus *j* for the child, mother and father respectively. We estimated the effects of parental educational attainment on each of the outcomes using the SNP-level associations with the mothers’ and fathers’ genotype and the outcomes and the association (coefficient and standard error) of each SNP and educational attainment from published GWAS[16]. We estimated the effect of parents’ education using inverse variance weighted (assuming no directional pleiotropy), weighted median (median estimate is unbiased), weighted mode (modal estimate is unbiased) estimators, and MR-Egger estimators (pleiotropy is independent of SNP-exposure association). These two-sample Mendelian randomization estimators make different assumptions about the structure of pleiotropy and would allow for estimation even in given specific forms of pleiotropy[52].

##### Multiple imputation

The questionnaire and, more rarely, administrative data had missing values. We addressed this using multiple imputation using Stata’s mi package. We included all covariates, exposures and outcomes in the imputation. The genetic variables, principal components, genotyping centre and chip for child, mother and father and the child’s sex, and birth year were included as regular variables as they had no missing data. Categorical variables with five or fewer categories were imputed using ordered logit, parental years of education and variables with more than five categories were imputed using truncated regression, test scores were imputed using predictive mean matching drawing from the ten nearest neighbours using a seed of 100, and truncated regression for the continuous or ordered categorical variables. We created twenty-five imputation datasets, and the analysis below was conducted across all datasets. We report the complete case analysis restricted to samples with no missing phenotypic data as a sensitivity analysis.

##### Sensitivity analyses

We investigated if our results differed by the child’s sex. We reran the imputation models in females and males and reran the phenotypic and Mendelian randomization analysis. We tested for differences between the estimates for females and males.

##### Non-imputed analysis

We repeated our primary analysis using the sample subset with no missing values. We estimated both the phenotypic association and within-family Mendelian randomization. Otherwise, all other parameters and methods were the same as the primary analysis.

##### Simulation

We investigated whether assortative mating could bias within-family Mendelian randomization estimates of dynastic effects using simulations. We simulated genotypes for one biallellic SNP for 40,000 mothers and fathers, and assorted the parents by genotype with a normally distributed error term using the following functions for mothers and fathers:

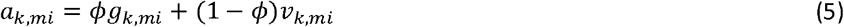

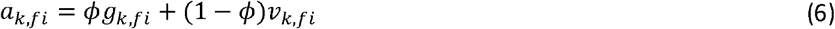

We then generated the children’s genotypes by randomly drawing alleles from the parents’ genotypes. Parents’ exposure was a function of their genotype and a standard normally distributed error term. Child phenotype was a function of parents’ phenotype (coefficient=1), and a standard normal error term. We assessed bias (i.e. deviations from one) for levels of assortment *ϕ* = {0,..,0.9}.

##### Software

We used Rstudio version 4.2.0, Stata version 17, Plink 2.00a2.3LM, and TwoSampleMR package version 0.5.6[53–56].

## Acknowledgements

This study includes data from the Norwegian Mother, Father and Child Cohort Study (MoBa) conducted by the Norwegian Institute of Public Health. The Norwegian Mother, Father and Child Cohort Study is supported by the Norwegian Ministry of Health and Care Services and the Ministry of Education and Research. We are grateful to all the participating families in Norway who take part in this on-going cohort study. We thank the Norwegian Institute of Public Health (NIPH) for generating high-quality genomic data. This research is part of the HARVEST collaboration, supported by the Research Council of Norway (#229624). We also thank the NORMENT Centre for providing genotype data (funded by the Research Council of Norway (#223273), South East Norway Health Authority and KG Jebsen Stiftelsen) in collaboration with deCODE Genetics. We further thank the Center for Diabetes Research, the University of Bergen for providing genotype data funded by the ERC AdG project SELECTionPREDISPOSED, Stiftelsen Kristian Gerhard Jebsen, Trond Mohn Foundation, the Research Council of Norway, the Novo Nordisk Foundation, the University of Bergen, and the Western Norway health Authorities (Helse Vest). This work was performed on the Tjeneste for Sensitive Data (TSD) facilities, owned by the University of Oslo, operated and developed by the TSD service group at the University of Oslo, IT-Department (USIT), using resources provided by Sigma2— the National Infrastructure for High Performance Computing and Data Storage in Norway (UNINETT).

## Contributions

AH, AMH and NMD were closely involved in conceptualising and designing the study. GDS and AH were involved in data and funding acquisition. NMD and AMH developed the pipeline. ECC, AMH, and NMD cleaned and ran the post-imputation QC on the MoBa data. NMD, AMH and AH performed data analyses. NMD wrote the first draft. AH and AMH played a key role in interpreting the results, planning additional analyses, and revising the manuscript. All authors contributed to and critically reviewed the manuscript.

## Data availability

The consent given by the participants does not allow for data storage on an individual level in repositories or journals. Researchers who want access to data sets for replication should apply to. Access to data sets requires approval from The Regional Committee for Medical and Health Research Ethics in Norway and an agreement with MoBa.

## Code availability

All the code used to clean and analyse the data for this study will be made available here https://github.com/nmdavies/moba_parent_education.

## References

1 Heckman JJ. Skill Formation and the Economics of Investing in Disadvantaged Children. Science 2006;312:1900–2. doi:10.1126/science.1128898

2 Morris TT, Hinke S, Pike L, et al. Implications of the genomic revolution for education research and policy. British Educational Res J 2022;:berj.3784. doi:10.1002/berj.3784

3 Young AI, Benonisdottir S, Przeworski M, et al. Deconstructing the sources of genotype-phenotype associations in humans. Science 2019;365:1396–400. doi:10.1126/science.aax3710

4 Branigan AR, McCallum KJ, Freese J. Variation in the Heritability of Educational Attainment: An International Meta-Analysis. Social Forces 2013;92:109–40. doi:10.1093/sf/sot076

5 Lee JJ, 23andMe Research Team, COGENT (Cognitive Genomics Consortium), et al. Gene discovery and polygenic prediction from a genome-wide association study of educational attainment in 1.1 million individuals. Nature Genetics 2018;50:1112–21. doi:10.1038/s41588-018-0147-3

6 Kong A, Thorleifsson G, Frigge ML, et al. The nature of nurture: Effects of parental genotypes. Science 2018;359:424–8. doi:10.1126/science.aan6877

7 Wang B, Baldwin JR, Schoeler T, et al. Robust genetic nurture effects on education: A systematic review and meta-analysis based on 38,654 families across 8 cohorts. The American Journal of Human Genetics 2021;108:1780–91. doi:10.1016/j.ajhg.2021.07.010

8 Brumpton B, Sanderson E, Heilbron K, et al. Avoiding dynastic, assortative mating, and population stratification biases in Mendelian randomization through within-family analyses. Nat Commun 2020;11:3519. doi:10.1038/s41467-020-17117-4

9 Howe LJ, Nivard MG, Morris TT, et al. Within-sibship GWAS improve estimates of direct genetic effects. Genetics 2021. doi:10.1101/2021.03.05.433935

10 Nivard MG, Belsky D, Harden KP, et al. Neither nature nor nurture: Using extended pedigree data to elucidate the origins of indirect genetic effects on offspring educational outcomes. PsyArXiv 2022. doi:10.31234/osf.io/bhpm5

11 Davies NM, Howe LJ, Brumpton B, et al. Within family Mendelian randomization studies. Human Molecular Genetics 2019;28:R170–9. doi:10.1093/hmg/ddz204

12 Davey Smith G, Ebrahim S. ‘Mendelian randomization’: can genetic epidemiology contribute to understanding environmental determinants of disease? Int J Epidemiol 2003;32:1–22. doi:10.1093/ije/dyg070

13 Sanderson E, Glymour MM, Holmes MV, et al. Mendelian randomization. Nat Rev Methods Primers 2022;2:6. doi:10.1038/s43586-021-00092-5

14 Davies NM, Holmes MV, Davey Smith G. Reading Mendelian randomisation studies: a guide, glossary, and checklist for clinicians. BMJ 2018;:k601. doi:10.1136/bmj.k601

15 Katan M. Apoupoprotein E Isoforms, Serum Cholesterol, And Cancer. The Lancet 1986;327:507–8. doi:10.1016/S0140-6736(86)92972-7

16 Okbay A, Wu Y, Wang N, et al. Polygenic prediction of educational attainment within and between families from genome-wide association analyses in 3 million individuals. Nat Genet 2022;54:437–49. doi:10.1038/s41588-022-01016-z

17 Sanderson E, Davey Smith G, Windmeijer F, et al. An examination of multivariable Mendelian randomization in the single-sample and two-sample summary data settings. Int J Epidemiol Published Online First: 10 December 2018. doi:10.1093/ije/dyy262

18 Mathieson I, McVean G. Differential confounding of rare and common variants in spatially structured populations. Nat Genet 2012;44:243–6. doi:10.1038/ng.1074

19 Skrivankova VW, Richmond RC, Woolf BAR, et al. Strengthening the reporting of observational studies in epidemiology using mendelian randomisation (STROBE-MR): explanation and elaboration. BMJ 2021;:n2233. doi:10.1136/bmj.n2233

20 Statistics | Eurostat. https://ec.europa.eu/eurostat/databrowser/view/EDAT_LFSE_04__custom_4612872/default/table?lang=en (accessed 20 Jan 2023).

21 Sanderson E, Windmeijer F. A weak instrument F-test in linear IV models with multiple endogenous variables. Journal of Econometrics Published Online First: June 2015. doi:10.1016/j.jeconom.2015.06.004

22 Sadreev II, Elsworth BL, Mitchell RE, et al. Navigating sample overlap, winner’s curse and weak instrument bias in Mendelian randomization studies using the UK Biobank. Epidemiology 2021. doi:10.1101/2021.06.28.21259622

23 Belsky J, Conger R, Capaldi DM. The intergenerational transmission of parenting: Introduction to the special section. Developmental Psychology 2009;45:1201–4. doi:10.1037/a0016245

24 Ahlburg D. Intergenerational transmission of health. The American Economic Review 1998;88:265–70.

25 Black SE, Devereux PJ, Salvanes KG. Why the Apple Doesn’t Fall Far: Understanding Intergenerational Transmission of Human Capital. American Economic Review 2005;95:437–49. doi:10.1257/0002828053828635

26 Blanden J, Gregg P, Macmillan L. Accounting for Intergenerational Income Persistence: Noncognitive Skills, Ability and Education. The Economic Journal 2007;117:C43–60. doi:10.1111/j.1468-0297.2007.02034.x

27 Hjalmarsson R, Lindquist MJ. The origins of intergenerational associations in crime: Lessons from Swedish adoption data. Labour Economics 2013;20:68–81. doi:10.1016/j.labeco.2012.11.001

28 Plomin R, Bergeman CS. The nature of nurture: Genetic influence on “environmental” measures. Behavioral and Brain Sciences 1991;14:373–86. doi:10.1017/S0140525X00070278

29 Plomin R, Shakeshaft NG, McMillan A, et al. Nature, nurture, and expertise. Intelligence 2014;45:46–59. doi:10.1016/j.intell.2013.06.008

30 Turkheimer E, Haley A, Waldron M, et al. Socioeconomic status modifies heritability of IQ in young children. Psychological Science 2003;14:623.

31 Torvik FA, Eilertsen EM, McAdams TA, et al. Mechanisms linking parental educational attainment with child ADHD, depression, and academic problems: a study of extended families in The Norwegian Mother, Father and Child Cohort Study. J Child Psychol Psychiatry 2020;61:1009–18. doi:10.1111/jcpp.13197

32 Cheesman R, Eilertsen EM, Ahmadzadeh YI, et al. How important are parents in the development of child anxiety and depression? A genomic analysis of parent-offspring trios in the Norwegian Mother Father and Child Cohort Study (MoBa). BMC Med 2020;18:284. doi:10.1186/s12916-020-01760-1

33 Isungset MA, Conley D, Zachrisson HD, et al. Social and genetic associations with educational performance in a Scandinavian welfare state. Proc Natl Acad Sci USA 2022;119:e2201869119. doi:10.1073/pnas.2201869119

34 Wertz J, Moffitt TE, Agnew-Blais J, et al. Using DNA From Mothers and Children to Study Parental Investment in Children’s Educational Attainment. Child Dev 2020;91:1745–61. doi:10.1111/cdev.13329

35 Chan TW, Boliver V. The Grandparents Effect in Social Mobility: Evidence from British Birth Cohort Studies. Am Sociol Rev 2013;78:662–78. doi:10.1177/0003122413489130

36 Fewell Z, Davey Smith G, Sterne JAC. The Impact of Residual and Unmeasured Confounding in Epidemiologic Studies: A Simulation Study. American Journal of Epidemiology 2007;166:646–55. doi:10.1093/aje/kwm165

37 Macmillan L, Tominey E. Parental inputs and socio-economic gaps in early child development. J Popul Econ Published Online First: 17 August 2022. doi:10.1007/s00148-022-00917-x

38 Kalil A, Mogstad M, Rege M, et al. Father Presence and the Intergenerational Transmission of Educational Attainment. Journal of Human Resources 2016;51:869–99. doi:10.3368/jhr.51.4.1014-6678R

39 Gould ED, Simhon A, Weinberg BA. Does Parental Quality Matter? Evidence on the Transmission of Human Capital Using Variation in Parental Influence from Death, Divorce, and Family Size. Journal of Labor Economics 2020;38:569–610. doi:10.1086/705904

40 Bergero R, Ellis P, Haerty W, et al. Meiosis and beyond – understanding the mechanistic and evolutionary processes shaping the germline genome. Biol Rev 2021;96:822–41. doi:10.1111/brv.12680

41 Sanderson E, Richardson TG, Morris TT, et al. Estimation of causal effects of a time-varying exposure at multiple time points through multivariable mendelian randomization. PLoS Genet 2022;18:e1010290. doi:10.1371/journal.pgen.1010290

42 Morris TT, Heron J, Sanderson ECM, et al. Interpretation of Mendelian randomization using a single measure of an exposure that varies over time. International Journal of Epidemiology 2022;51:1899–909. doi:10.1093/ije/dyac136

43 Demange PA, Hottenga JJ, Abdellaoui A, et al. Estimating effects of parents’ cognitive and non-cognitive skills on offspring education using polygenic scores. Nat Commun 2022;13:4801. doi:10.1038/s41467-022-32003-x

44 Balbona JV, Kim Y, Keller MC. Estimation of Parental Effects Using Polygenic Scores. Behav Genet 2021;51:264–78. doi:10.1007/s10519-020-10032-w

45 Tudball MJ, Smith GD, Zhao Q. Almost exact Mendelian randomization. Published Online First: 2022. doi:10.48550/ARXIV.2208.14035

46 Magnus P, Birke C, Vejrup K, et al. Cohort Profile Update: The Norwegian Mother and Child Cohort Study (MoBa). Int J Epidemiol 2016;45:382–8. doi:10.1093/ije/dyw029

47 Paltiel L, Anita H, Skjerden T, et al. The biobank of the Norwegian Mother and Child Cohort Study – present status. Nor J Epidemiol 2014;24. doi:10.5324/nje.v24i1-2.1755

48 Sandsør AMJ, Zachrisson HD, Karoly LA, et al. The Widening Achievement Gap Between Rich and Poor in a Nordic Country. Educational Researcher 2023;:0013189X2211425. doi:10.3102/0013189X221142596

49 Corfield EC, Frei O, Shadrin AA, et al. The Norwegian Mother, Father, and Child cohort study (MoBa) genotyping data resource: MoBaPsychGen pipeline v.1. Genetics 2022. doi:10.1101/2022.06.23.496289

50 Helgeland Ø, Vaudel M, Sole-Navais P, et al. Characterization of the genetic architecture of infant and early childhood body mass index. Nat Metab 2022;4:344–58. doi:10.1038/s42255-022-00549-1

51 Helgeland Ø, Vaudel M, Juliusson PB, et al. Genome-wide association study reveals dynamic role of genetic variation in infant and early childhood growth. Nat Commun 2019;10:4448. doi:10.1038/s41467-019-12308-0

52 Hemani G, Bowden J, Davey Smith G. Evaluating the potential role of pleiotropy in Mendelian randomization studies. Human Molecular Genetics 2018;27:R195–208. doi:10.1093/hmg/ddy163

53 StataCorp. Stata Statistical Software: Release 17. 2021.

54 R Core Team. R: A Language and Environment for Statistical Computing. Vienna, Austria: : R Foundation for Statistical Computing 2017. https://www.R-project.org/

55 Hemani G, Zheng J, Elsworth B, et al. The MR-Base platform supports systematic causal inference across the human phenome. eLife 2018;7. doi:10.7554/eLife.34408

56 Purcell S, Neale B, Todd-Brown K, et al. PLINK: a tool set for whole-genome association and population-based linkage analyses. Am J Hum Genet 2007;81:559–75. doi:10.1086/519795

