## Supplementary Figures for "Intergenerational effects of parental educational attainment on parenting and childhood educational outcomes: Evidence from MoBa using within-family Mendelian randomization"

**Supplementary materials**

#### **Supplementary Figure 1: STROBE Flow chart of inclusion and exclusion from the study**

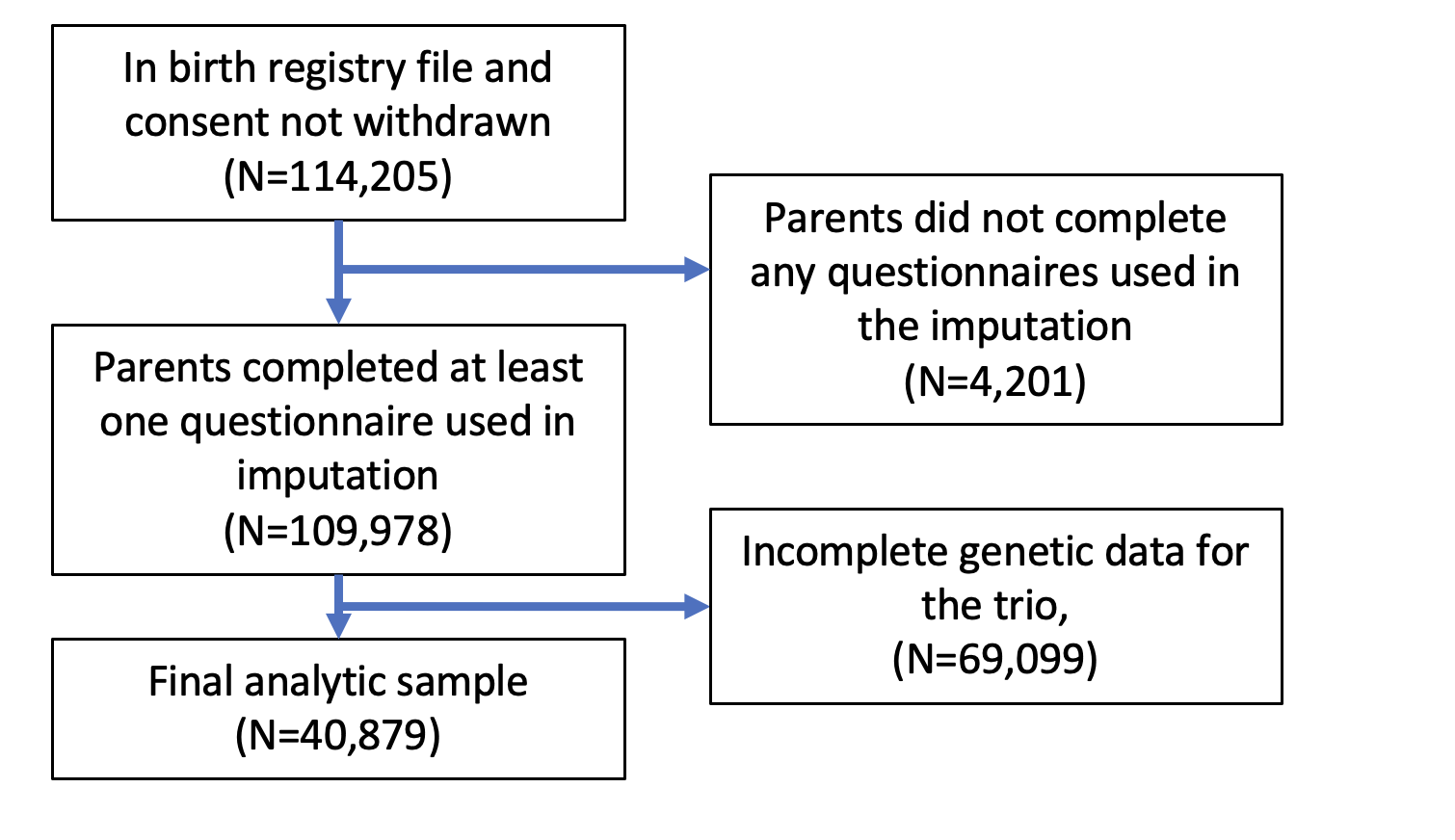

#### **Supplementary Figure 2: Sex stratified FEMALES Effect of parents’ educational attainment on children’s nationally standardised test scores, estimated using multivariable-adjusted regression (OLS) and within-family Mendelian randomization (WF), estimated on the full sample using multiple imputation (N=20,866). Mean difference in standard deviations and 95% confidence intervals in test scores per year of parental education reported.**

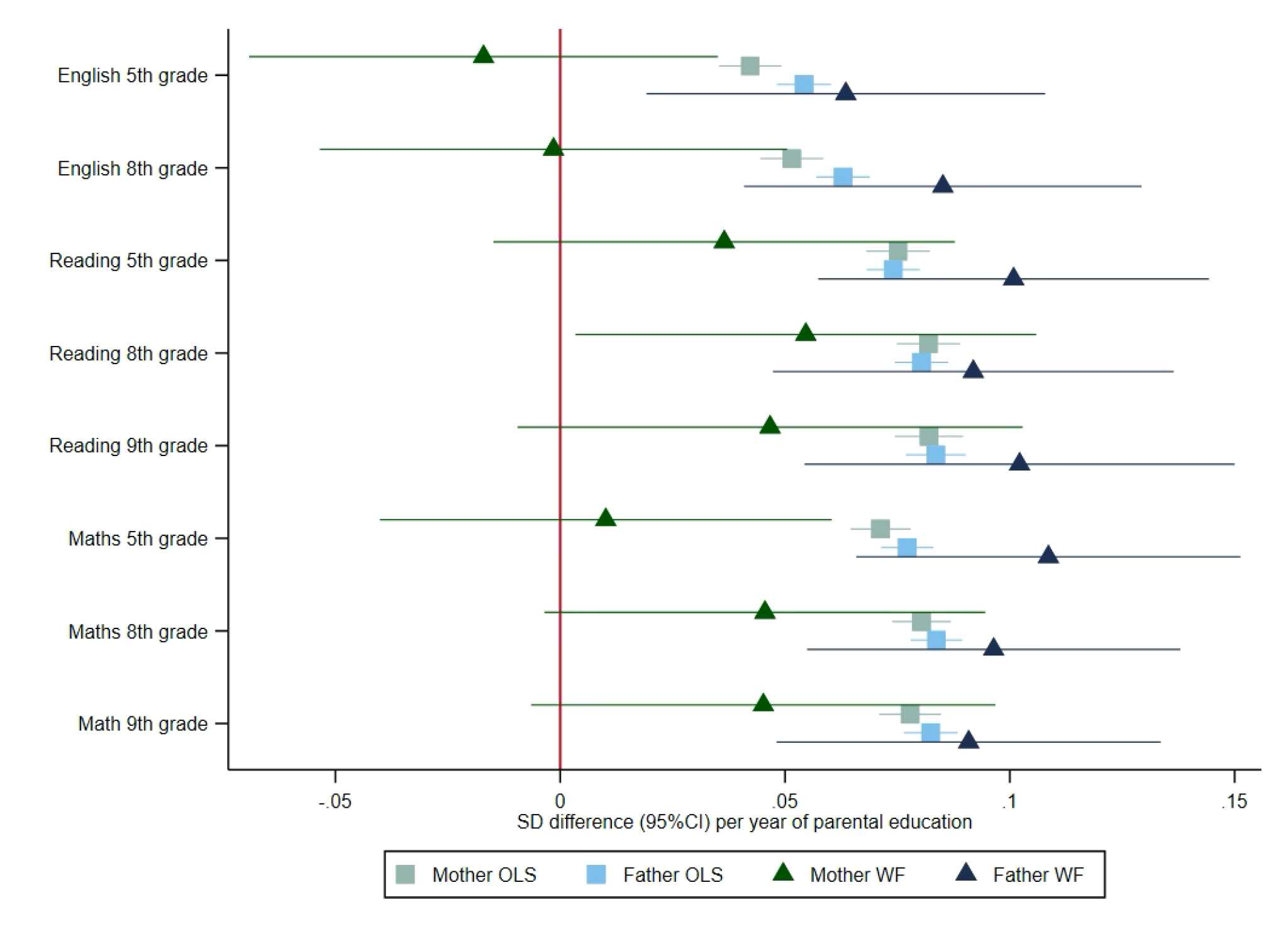

#### **Supplementary Figure 2: Sex stratified MALES Effect of parents’ educational attainment on children’s nationally standardised test scores, estimated using multivariable-adjusted regression (OLS) and within-family Mendelian randomization (WF), estimated on the full sample using multiple imputation (N=20,013). Mean difference in standard deviations and 95% confidence intervals in test scores per year of parental education reported.**

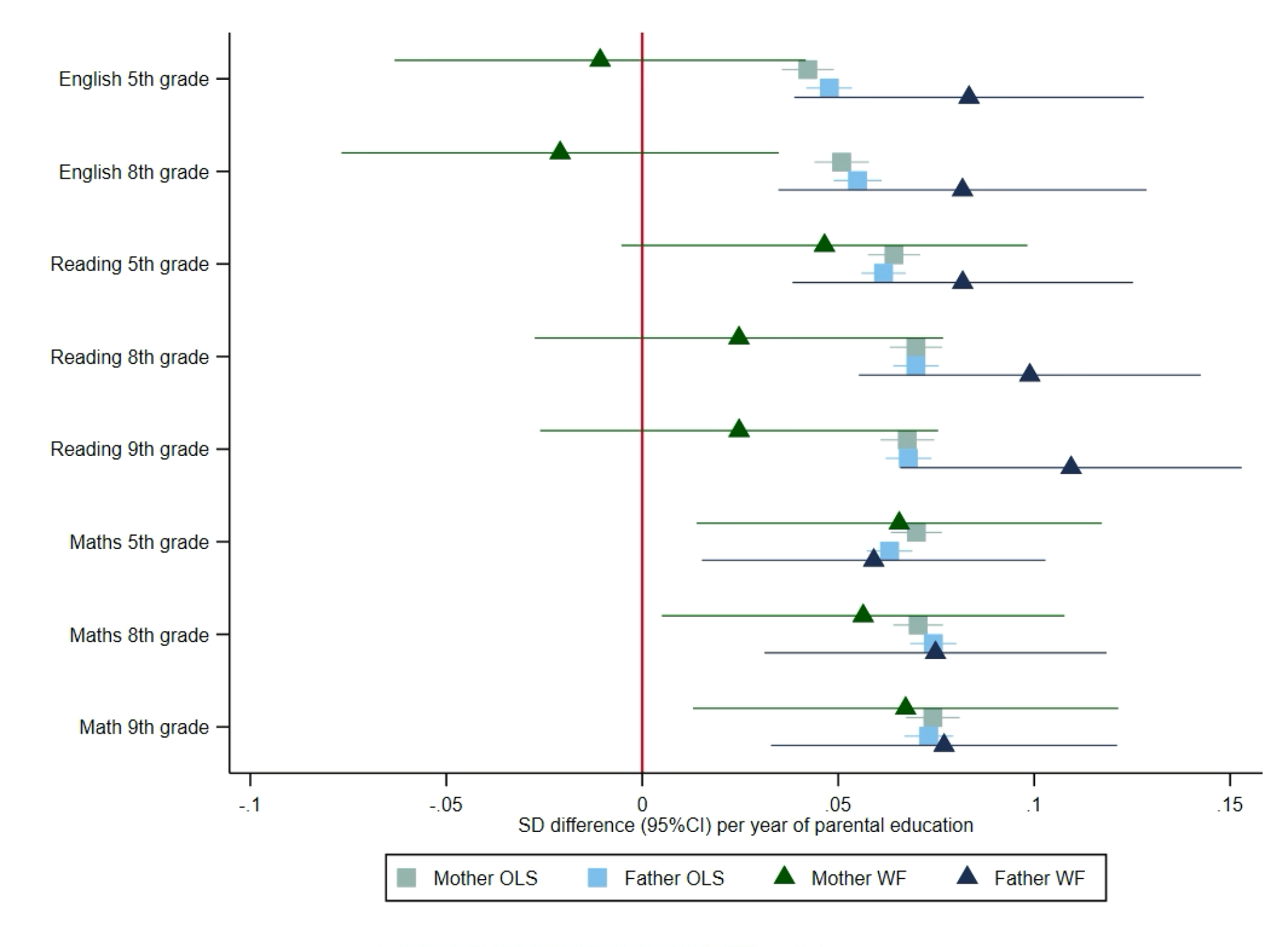

#### **Supplementary Figure 3: Sex-stratified FEMALES Effect of parents’ educational attainment on questionnaire measures of reading and communication skills and parental nurturing at age 5, estimated using multivariable-adjusted regression (OLS) and within-family Mendelian randomization (WF), estimated on the full sample using multiple imputation (N=20,866).**

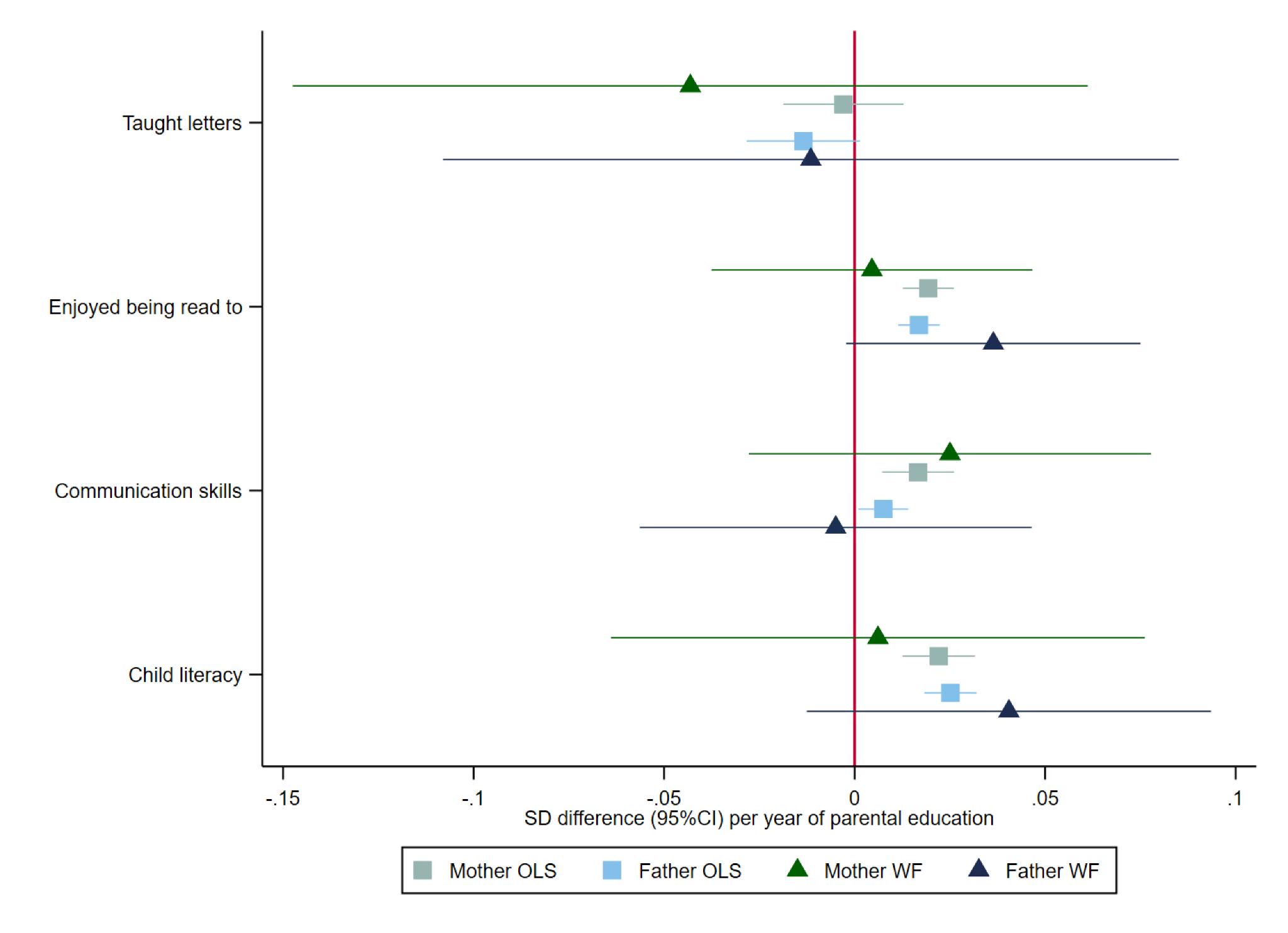

#### **Supplementary Figure 4: Sex-stratified MALES Effect of parents’ educational attainment on questionnaire measures of reading and communication skills and parental nurturing at age 5, estimated using multivariable-adjusted regression (OLS) and within-family Mendelian randomization (WF), estimated on the full sample using multiple imputation (N=20,021).**

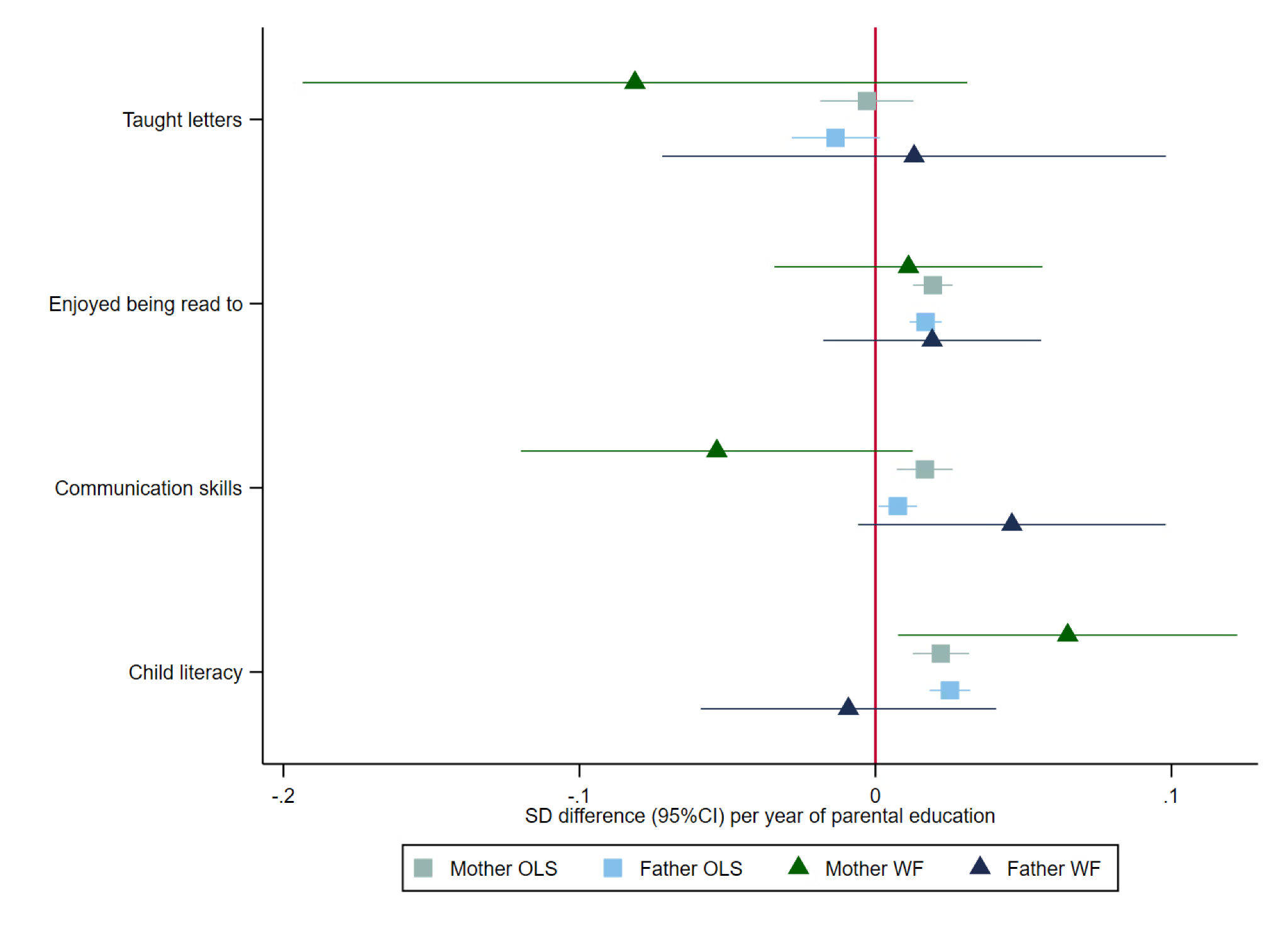

#### **Supplementary Figure 6: Sex stratified FEMALES Effect of parents’ educational attainment on questionnaire measures of early educational attainment and parental nurturing at age 8, estimated using multivariable-adjusted regression (OLS) and within-family Mendelian randomization (WF), estimated on the full sample using multiple imputation (N=20,866).**

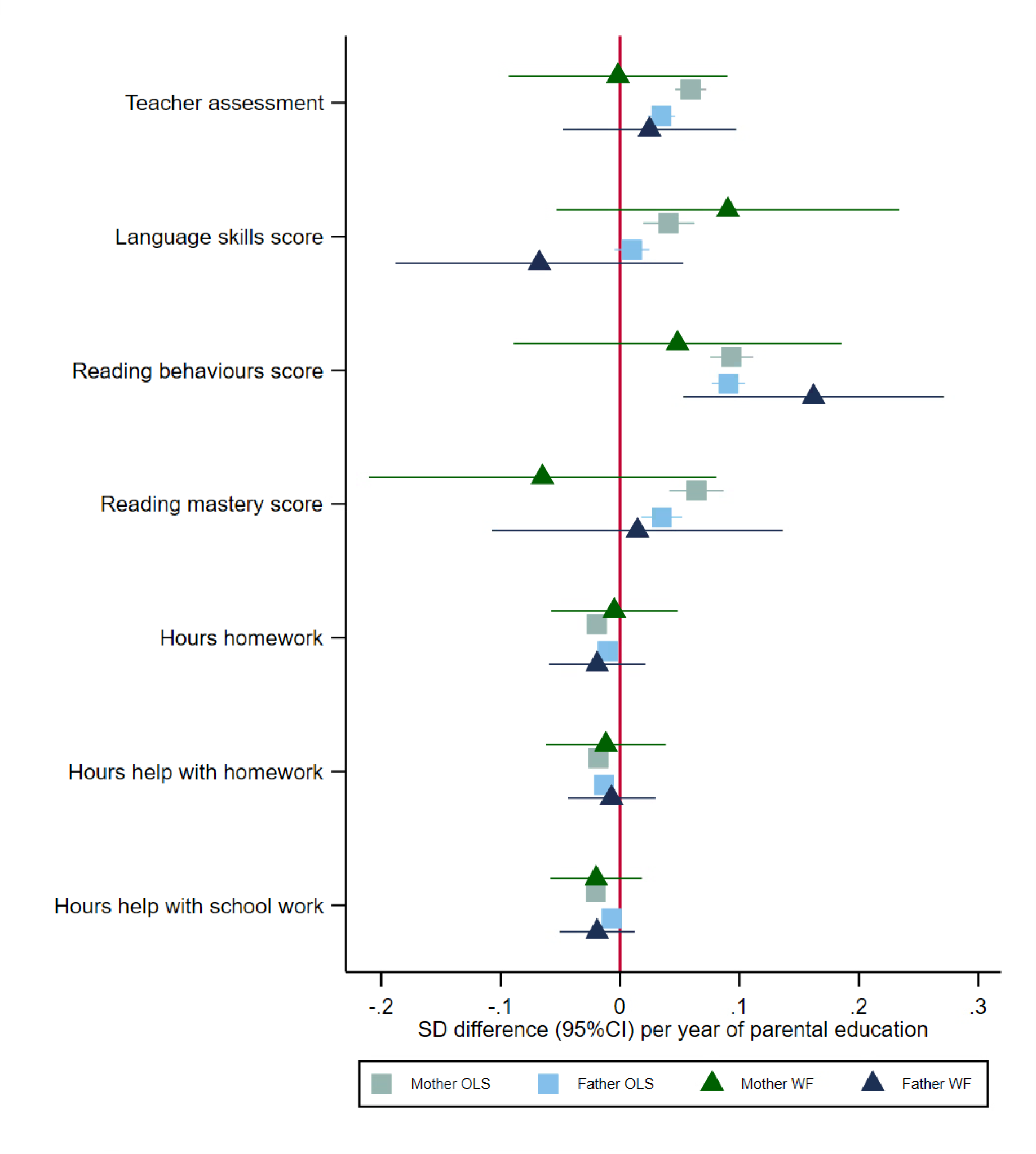

#### **Supplementary Figure 7: Sex stratified MALES Effect of parents’ educational attainment on questionnaire measures of early educational attainment and parental nurturing at age 8, estimated using multivariable-adjusted regression (OLS) and within-family Mendelian randomization (WF), estimated on the full sample using multiple imputation (N=20,021).**

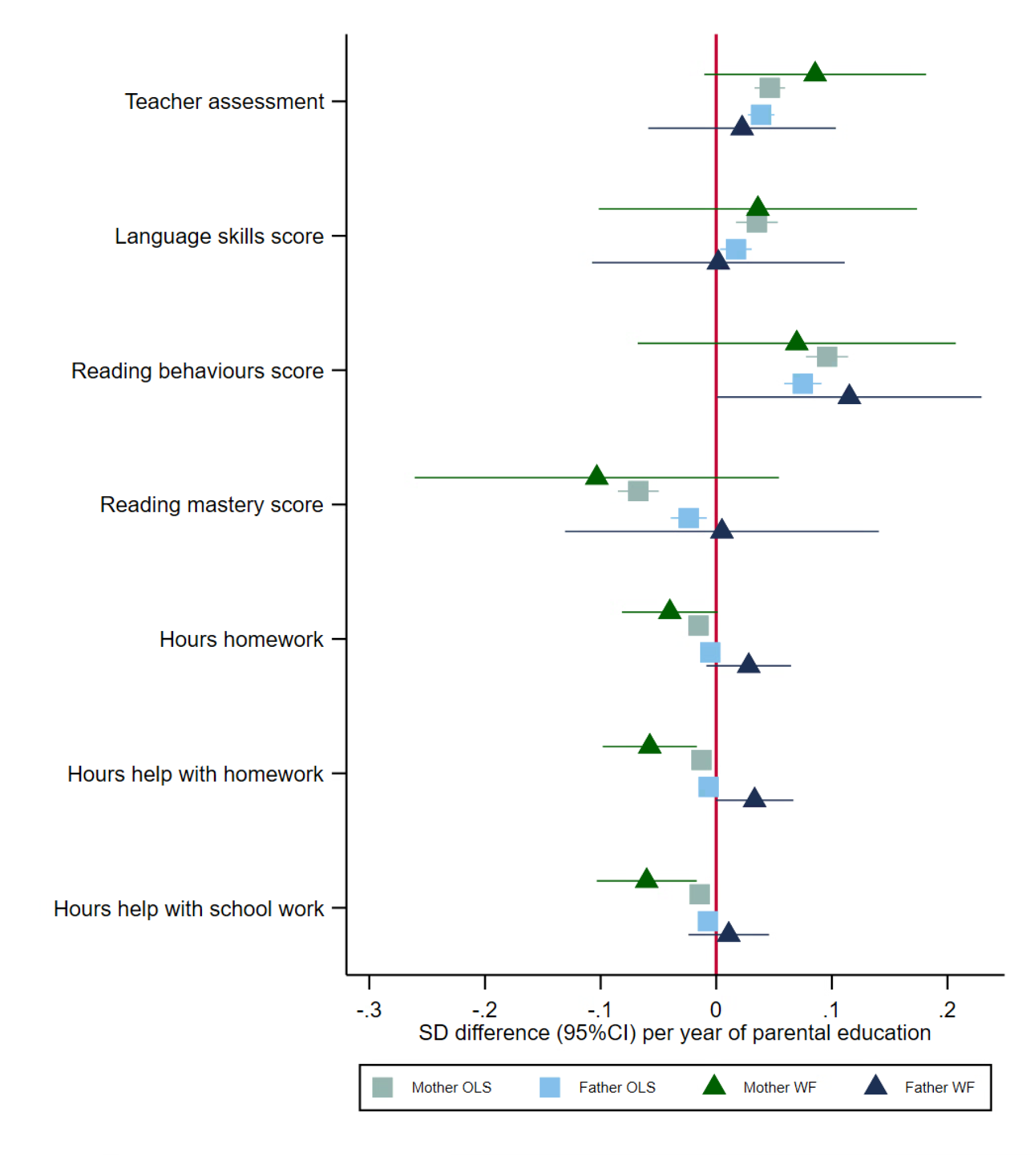

#### **Supplementary Figure 8: Simulation comparing ordinary least squares (OLS), standard Mendelian randomization and within-family Mendelian randomization estimates of the effect of mothers on their children. The true effect size of the dynastic effect is one, indicated by the red line. Within-family Mendelian randomization estimators can provide an unbiased estimate of the effects of parents on their offspring, even in the presence of assortative mating. At very high levels of assortment, the precision of these estimators falls. OLS and standard Mendelian randomization are biased.**

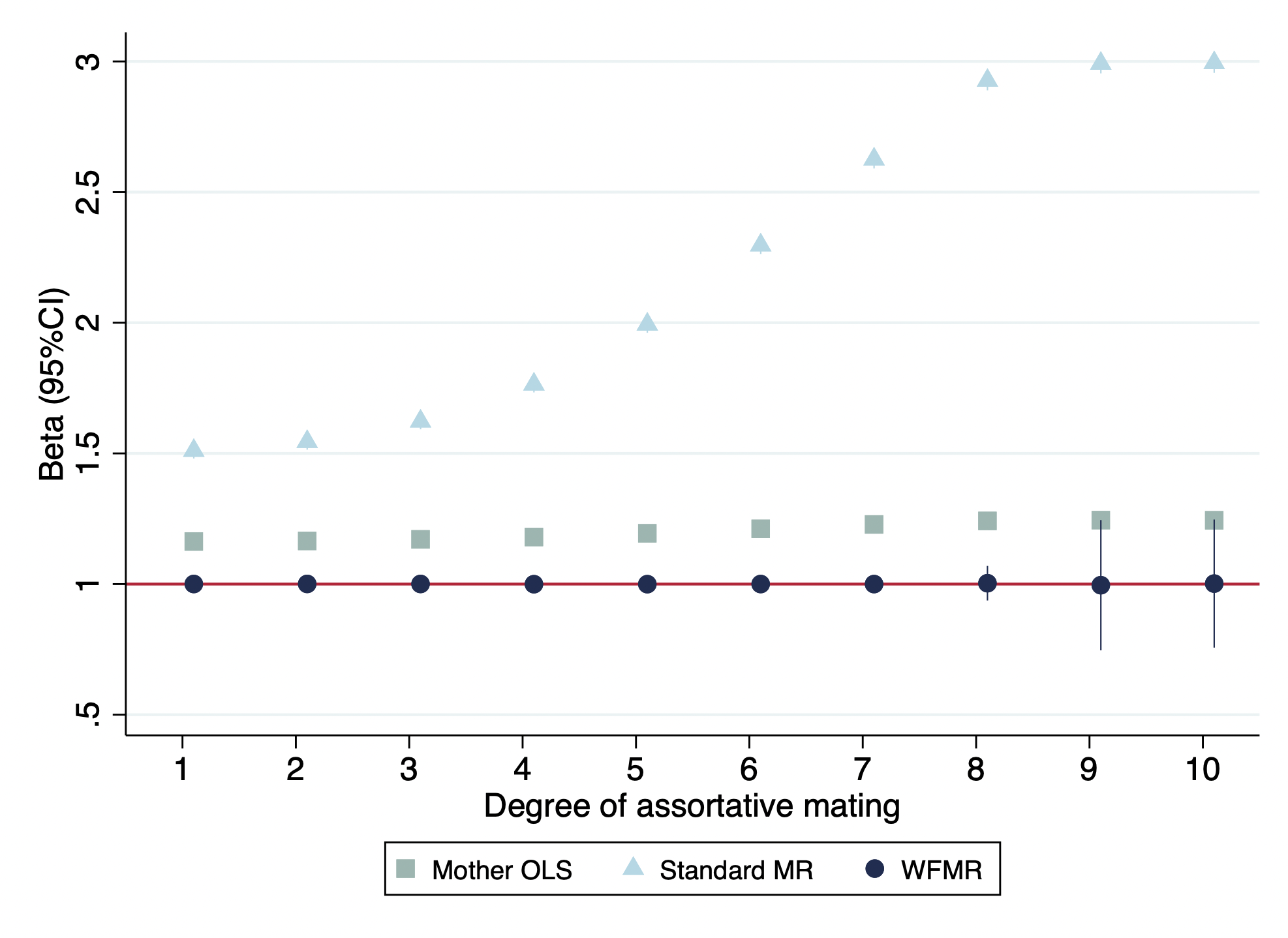
